# Angular changes in implants placed in the anterior maxillae of adults: A cephalometric pilot study

**DOI:** 10.1101/2020.03.23.20041855

**Authors:** Balazs Feher, Reinhard Gruber, Andre Gahleitner, Ales Celar, Philipp Luciano Necsea, Christian Ulm, Ulrike Kuchler

## Abstract

**Objectives:** Completion of adolescent growth represents the earliest time point for implant placement, yet craniofacial growth persists into adulthood and may affect implant position. We aimed to assess whether implants placed in the anterior maxillae of adults show angular changes.

**Methods:** We conducted a cephalometric pilot study in postpubertal patients with no growth disorders, skeletal malformations, or parafunctions. The patients received a single implant in the anterior maxilla and no orthodontic or orthognathic treatment afterwards. We measured angular changes on cephalograms taken immediately and at least 5 years postoperatively in a standardized setting.

**Results:** In a total of 21 patients (30.2 ± 11.5 years at surgery) after a mean follow-up time of 8.6 ± 1.3 years, 62% of implants showed counterclockwise rotations (1.8 ± 1.0 degrees) and 19% of implants showed clockwise rotations (2.4 ± 1.1 degrees). Angular changes were more frequent in males (100% *vs*. 58%) and patients under 30 at surgery (85% *vs*. 63%). Mean absolute differences were larger in males (1.8 ± 1.0 degrees *vs*. 1.3 ± 1.4 degrees) and patients under 30 at surgery (1.5 ± 1.4 degrees *vs*. 1.1 ± 1.4 degrees). Regression analysis did not identify explanatory factors for the observed changes.

**Conclusions:** Implants placed in the anterior maxillae of adults show modest angular changes over time.

**Clinical relevance:** Changes in implant angles have potential functional and esthetic consequences.

## Introduction

Osseointegration is a ‘functional ankylosis’ similar to that observed in teeth following injuries^1,2^. Dental implants, like ankylosed teeth, do not follow the growth of the alveolar processes during eruption^3^. The placement of implants in growing jawbones thus results in the submersion of the implants over time, relative to adjacent erupting teeth^4^. In order to avoid this phenomenon, the earliest time point recommended for implant placement is following the end of adolescence, when growth is thought to be completed^5,6^. However, craniofacial growth persists into adulthood^6,7^, causing significant dimensional changes of the facial skeleton in the long term^8-10^. With regard to dental implants, the clinical implications of this residual growth have largely been underestimated^11,12^. Previous work has suggested vertical changes in implants as a result of continuous growth in adults^13,14^. Recent data have given support to these findings and even pointed to a potentially high prevalence of infraocclusion^15-18^. Observed in adults, the described changes could not be explained by adolescent growth.

Humans are among the few species in which an adolescent growth spurt can be observed^19^. This period of significant increase in height and weight^20^ triggers major changes in the jawbones^21^. Implant therapy during adolescence is restricted to cases of extensive hypodontia^22^. Adolescence ends with the closing of the epiphyses of long bones, typically around 18 years of age in males and 15 in females^23^. Some surgical protocols consider individual variability in aging and thus recommend a more conservative approach of placing implants at a slightly higher age^24^. With most of the skeletal growth completed by the end of puberty, implant placement starting at early adulthood is generally considered safe. Nevertheless, findings on the effects of continuous craniofacial growth have raised the question whether clinically relevant changes still can occur in the adult patient.

Compared with previous work describing vertical changes^13-18^, data on possible angular changes in implants due to residual craniofacial growth are lacking. Understanding potential angular changes in implants is important for multiple reasons. In the anterior maxilla, the palatal crown surfaces of incisors guide protrusion and canines play an important role in guiding laterotrusion^25^; changes in implant angles could lead functional issues. Moreover, implant crown esthetics are essential for clinical success^26,27^. It is apparent that in an exposed area such as the anterior maxilla, angular changes could undermine optimal results. Cephalometry is a routine radiographic tool used in orthodontics and orthognathic surgery. Structures of the head skeleton as well as their spatial relationships can be accurately measured using cephalometry^28^. To understand the possible effect of residual craniofacial growth, we applied cephalometry in this pilot study to measure long-term angular changes in implants in the anterior maxillae of adult patients.

## Methods

### Experimental design

We conducted a long-term cephalometric pilot study that was designed in accordance with the Declaration of Helsinki. This study was conducted at a single center, the Medical University of Vienna, University Clinic of Dentistry. The study protocol was approved by the ethics committee of the Medical University of Vienna (*No*. 2174/2018). All recruited patients were fully informed about the procedure, the materials to be used in this study, their estimated exposure to radiation, the benefits, as well as potential risks and complications stemming from their participation in this study. All patients gave their written consent prior to participation in this study.

### Inclusion and exclusion criteria

We included patients that i) received a single implant in the anterior maxilla (*i*.*e*., canine to canine), ii) were at least 18 years of age at surgery, and iii) received their implants at least 5 years prior to this study. We excluded patients with i) birth defects with or without skeletal malformations (*e*.*g*., cleidocranial dysplasia), ii) congenital growth disorders (*e*.*g*., congenital growth hormone deficiency), iii) parafunctions (*e*.*g*., tongue thrust), iv) traumatic injuries prior to implant therapy, v) complications relating to the implant (*e*.*g*., peri-implantitis, fracture), as well as vi) orthodontic therapy or vii) orthognathic surgery (*e*.*g*., Le Fort osteotomy) following implant placement.

### Cephalometry

We used postoperative lateral cephalograms as baseline and took one follow-up lateral cephalogram per patient. Both cephalograms were taken in the same setting and using the same parameters (75 kV, 32 mAs, 3.9 m source-to-mid-sagittal-plane distance). The cephalograms were precisely standardized prior to analysis using a raster graphics editor (Photoshop, Adobe, Mountain View, CA, USA). Facial growth type was determined using Björk’s sum of the saddle angle, articular angle, and gonial angle^29^. The implant axis was defined as the straight line connecting the implant shoulder to the implant apex. The Sella-Nasion line was designated as the reference structure due to its stability^30^. The implant angle was defined as the plane angle between the Sella-Nasion line and the orofacial implant axis facing in anterior direction (**Figure 1**). The primary end point of this study was any change in the implant angle between baseline and follow-up. All lateral cephalograms were evaluated by a single researcher (PLN). To further ensure accuracy, the researcher was calibrated by unknowingly evaluating 21% of the complete radiographic dataset twice. The duplicate measurements were then compared by a different researcher (BF). Based on the comparison of the duplicates, the intraclass correlation coefficient of the evaluator was 99%.

**Figure 1.**
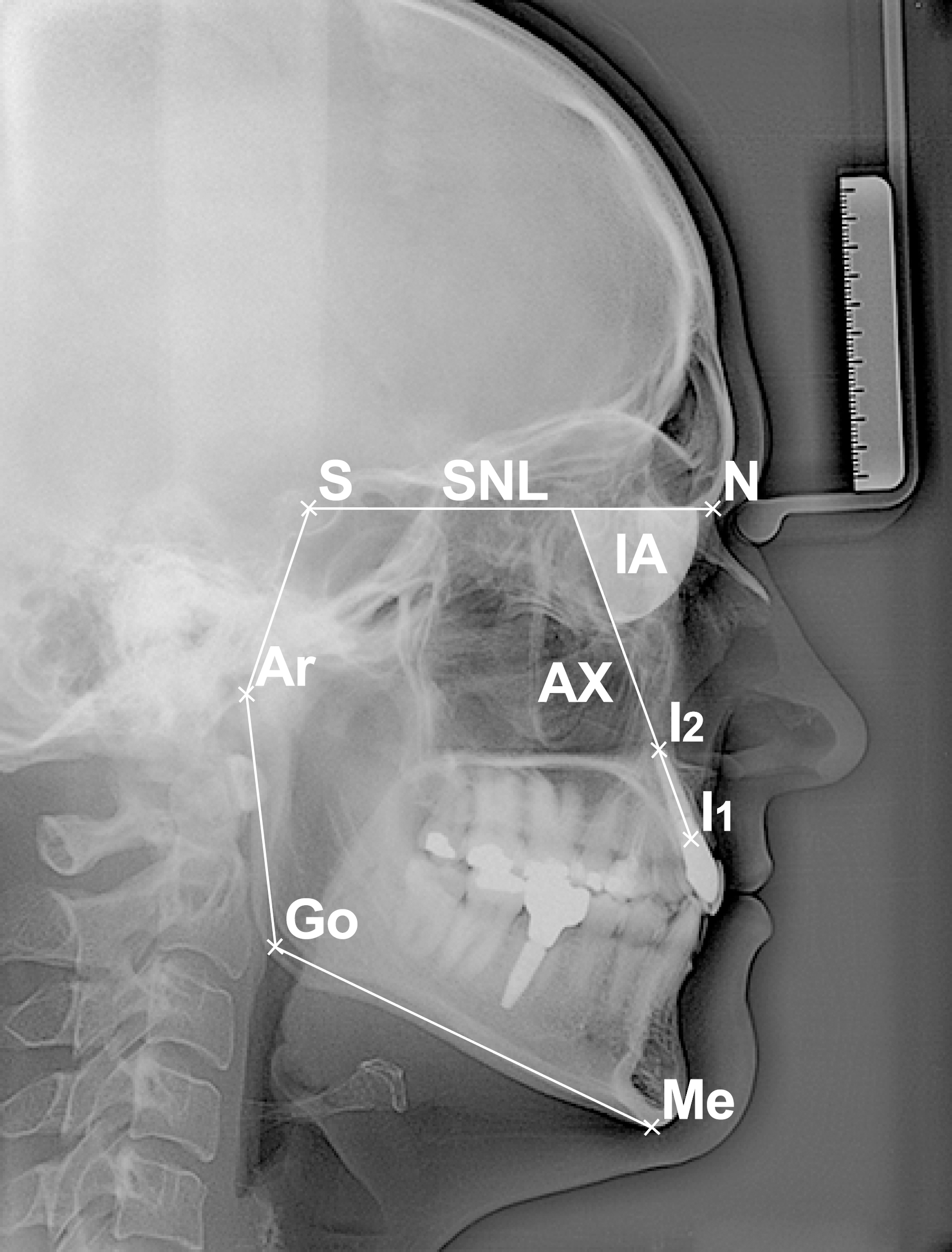
Radiographic parameters. Ar, Articulare; AX, orofacial implant axis; Go, Gonion; IA, implant angle; I1, implant shoulder; I2, implant apex; Me, Menton; N, Nasion; S, Sella; SNL, Sella-Nasion line.

### Statistics

Consistent with the pilot nature of this study, no sample size was calculated prior to patient enrollment, and statistical analyses were descriptive in nature. Data were first collected in a spreadsheet (Excel 16.29.1 for Mac, Microsoft Corporation, Redmond, WA, United States of America), error-proofed, and consequently analyzed using the R statistical computing environment (Version 3.6.1, R Core Team, Vienna, Austria). Descriptive statistical methods were used for subject characteristics and basic comparisons of subgroups (*e*.*g*., sex and age distribution). Mean values and standard deviations (SDs) were calculated for numerical variables. Kernel density estimates and histograms were used to visualize numerical variables (**Supplementary Figures 1a–c**). Linear regression analysis was further used to assess changes in the implant angles between baseline and follow-up. The implant angle at follow-up was set as the dependent variable, with the implant angle at baseline, sex, age, Björk’s angle sum, facial growth type, and follow-up time serving as independent variables.

## Results

### Study population

A total of 21 patients (mean age at follow-up: 38.9 ± 11.2 years, age range: 26–58 years, 57% female) completed the study after a mean follow-up time of 8.6 ± 1.3 years (range: 6.6–10.9 years). The patients’ mean age at surgery was 30.2 ± 11.5 years (range: 18–52 years). With regard to facial growth type, 67% of patients were brachyfacial (mean Björk’s sum: 385.3 ± 4.2 degrees, range: 376–390 degrees), 24% were mesofacial (mean Björk’s sum: 395.0 ± 2.1 degrees, range: 392–397 degrees), and 10% were dolichofacial (401.5 ± 0.7 degrees, range: 401–402 degrees). Subject characteristics are presented in **Table 1** and **Supplementary Figures 1a–c**.

**Table 1.** Subject characteristics

|  |  |
| --- | --- |
| **Total study population, n** | **21** |
| Sex, n (%) |  |
| Females | 12 (57.1) |
| Males | 9 (42.9) |
| Age, mean ± SD (range) in years | |
| At surgery | 30.2 ± 11.5 (18–52) |
| At follow-up | 38.9 ± 11.2 (26–58) |
| Growth type, n (%) |  |
| Brachyfacial | 14 (67) |
| Mesofacial | 5 (24) |
| Dolichofacial | 2 (10) |
| Follow-up, mean ± SD (range) in years | 8.6 ± 1.3 (6.6–10.9) |

SD, standard deviation.

### Changes in implant angles

To investigate angular changes in implants, we compared the implant angles on the baseline and follow-up radiographs. Angular changes were found in 81% of implants. A counterclockwise rotation (-^*ve*^ angular change) was found in 62% of implants (mean ± SD: −1.8 ± 1.0 degrees, range: −1 to −3 degrees). A clockwise rotation (+^*ve*^ angular change) was found in 19% of implants (mean ± SD: 2.4 ± 1.1 degrees, range: 1 to 4 degrees). No angular changes were found in 19% of implants. Angular changes were more frequent in males than in females (100% versus 58%). Mean absolute differences between baseline and follow-up were also larger in males than in females (1.8 ± 1.0 degrees versus 1.3 ± 1.4 degrees). Angular changes were more frequent in patients under 30 at surgery than patients at least 30 years old at baseline (85% versus 63%). Mean absolute differences between baseline and follow-up were slightly larger between patients under 30 and patients at least 30 years old at surgery (1.5 ± 1.4 degrees versus 1.1 ± 1.4 degrees). Together, these results indicate the possibility of changes in the angles of implants placed in the anterior maxillae of adults (**Table 2**).

**Table 2.** Changes in implant angles

|  | **Frequency,**<br>**%** | **Range,**<br>**degrees** | **Mean ± SD,**<br>**degrees** | **Abs. mean ± SD,**<br>**degrees** |
| --- | --- | --- | --- | --- |
| Sex |  |  |  |  |
| Females | 58 | -4 to 3 | -0.4 ± 1.8 | 1.3 ± 1.4 |
| Males | 100 | -3 to 3 | -0.9 ± 1.9 | 1.8 ± 1.0 |
| Age groups |  |  |  |  |
| Under 20 at surgery | 80 | -2 to 3 | 0.4 ± 2.1 | 1.6 ± 1.1 |
| Over 20 and under 30 at surgery | 88 | -4 to 1 | -1.3 ± 1.5 | 1.5 ± 1.2 |
| Over 30 and under 40 at surgery | 50 | -3 to 0 | -1.5 ± 2.1 | 1.5 ± 2.1 |
| Over 40 at surgery | 67 | -3 to 3 | -0.3 ± 2.0 | 1.3 ± 1.4 |
| Growth type |  |  |  |  |
| Brachyfacial | 79 | -4 to 3 | -0.4 ± 2.0 | 1.6 ± 1.2 |
| Mesofacial | 80 | -3 to 0 | -1.6 ± 1.3 | 1.6 ± 1.3 |
| Dolichofacial | 50 | 0 to 1 | 0.5 ± 0.7 | 0.5 ± 0.7 |

Positive angular changes represent counterclockwise rotations. Abs, absolute; SD, standard deviation.

### Linear regression analysis

To further analyze angular changes as well as determine whether demographic or growth-related factors could have an effect on them, we applied linear regression analysis in an explorative manner. The analysis returned a slope of regression of 0 (*p < 0*.*001*) (**Figure 2**). Further, none of the assessed predictors (implant angle at baseline, sex, age, Björk’s angle sum, facial growth type, and follow-up time) had an influence on the implant angle at follow-up.

**Figure 2.**
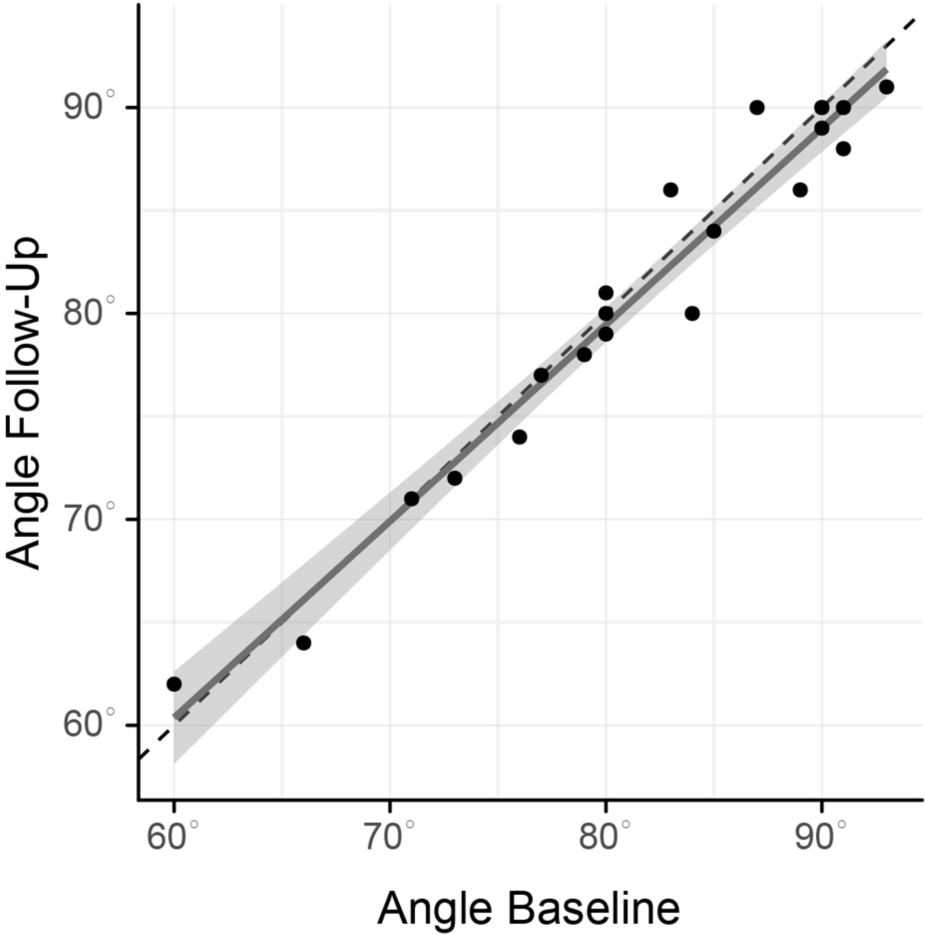
Linear regression analysis. The dashed line shows no change. The dark grey line represents the regression line. The light grey area surrounding the dark grey line represents the corresponding 95% confidence interval. Slope of regression = 0 (p < 0.001).

## Discussion

Evidence on long-term changes in implant positions in adult patients is accumulating. Following early data from preclinical models^4,31^ and clinical studies in adolescents^32^, findings from adults have substantiated the possibility of vertical changes in implant position, up to the point of infraocclusion^13-16^. The present pilot study is the first to report angular changes over the course of adulthood. Based on a cephalometric analysis, we found that after a mean follow-up time of 8.6 years, implants placed in the anterior maxillae showed modest rotational changes ranging from 3 degrees counterclockwise to 4 degrees clockwise. These findings are important for they show the possibility of angular changes in implants placed in adult patients. Our findings further relate to those of others as at 81%, the prevalence of angular changes in our patient cohort is comparable to that of vertical changes (73%) described recently^16^.

In our patient cohort, angular changes were found in 58% of females and 100% of males. The sex differences are in contrast to the findings of others as with just one exception^33^, previous work did not identify a predisposing role of sex on vertical changes in implants^13,17,18,34^. With regard to age at surgery, angular changes were found in 85% of patients under 30 and 63% of patients at least 30 years old at surgery. The differences between different age groups are in accordance with some previous findings^34^ and in contrast to others^13,16,17^. The gradual decline of growth over time could explain how age at surgery influences long-term changes in implants. However, the existing literature does not unequivocally back up that theory^13,16,17^. We further found that the patient cohort showed a high individual variety in the extent and directions of rotational changes. Consistent with the pilot nature of this study, we did not conduct tests for statistical significance. In order to test the significance of angular changes or evaluate potential predictors, the threshold for a clinically relevant angular change has to be defined first by the scientific community. To assist future research into this area, we calculated sample sizes for theoretical thresholds of 1 to 7 degrees (**Supplementary Table 1**).

Computer-aided standardization prior to analysis as well as blinded observer calibration helped ensure precision and limit measurement error. Cephalometric analysis is inherently observer dependent. Intra-observer variability thus has to be minimized prior to analysis. Particular attention was given to the consistent marking of the Sella as it is a ‘floating’ landmark; all other landmarks relevant to the analysis of angular changes are discrete structures (*i*.*e*., nasofrontal suture, implant body). We thus believe the angular changes measured are not due to measurement error. The angular changes described herein ranging from 3 degrees counterclockwise to 4 degrees clockwise are not so substantial as to prevent implant placement starting at early adulthood. Nevertheless, the data give support to previous work^16-18^ highlighting the relevance of continuous craniofacial growth in implant dentistry. It remains open at what threshold angular changes become relevant to the patient; the present study did not evaluate that as the possibility of angular changes first had to be confirmed. While vertical changes are noticed by over 60% of affected patients, they are not necessarily dissatisfied as a result^16,18^. Nevertheless, the esthetics of implant restorations in the anterior maxilla are highly relevant to patients. It is thus reasonable to assume that angular changes in the esthetic zone could cause a high degree of dissatisfaction.

Limitations of the present pilot study include its retrospective design, its relatively small sample size, as well as its reliance on two-dimensional radiographic imaging. Alternatives to lateral cephalometry include three-dimensional cone beam computer tomography. However, metal streak artifacts associated with computer tomography could make it difficult to accurately measure implant angles. Three-dimensional magnetic resonance cephalometry^35^ could be utilized to overcome the potential limitations associated with the use of lateral cephalograms^36,37^. The increasing amount of data supporting changes in implants placed in adults underscores the importance of future research into this field. Further studies with a prospective study design could take advantage of higher sample sizes and three-dimensional cephalometry to better evaluate angular implant changes in the anterior maxillae of adults. In the present study, regression analysis failed to identify significant explanatory factors for the observed changes in implant angles. Nevertheless, the findings should be considered relevant and basically favorable because while we showed that angular changes can occur in implants over the course of adulthood, their scale does not indicate that we should reconsider implant therapy starting at early adulthood.

## Conclusions

Within the limitations of this pilot study, it can be concluded that implants placed in the anterior maxillae of adult patients show modest angular changes in the long term.

## Data Availability

All data referred to in the manuscript are available from the corresponding author upon reasonable request.

## Acknowledgements

The content is solely the responsibility of the authors and does not necessarily represent the official views of the Medical University of Vienna. The authors thank Prof. Ales Celar for his advice on cephalometry and Mr. Stefan Lettner for the statistical analysis. All authors agree to be accountable for all aspects of the work. The present study did not receive any funding.

---

**Supplementary Table 1.** Sample size calculation

| Change in implant angle, degrees | Sample size, n<br>Power = 0.80 | Sample size, n<br>Power = 0.90 | Sample size, n<br>Power = 0.95 | Sample size, n<br>Power = 0.99 |
| --- | --- | --- | --- | --- |
| 1 | 494 | 660 | 815 | 1152 |
| 2 | 125 | 167 | 206 | 290 |
| 3 | 57 | 75 | 93 | 130 |
| 4 | 33 | 44 | 53 | 74 |
| 5 | 22 | 29 | 35 | 48 |
| 6 | 16 | 21 | 25 | 34 |
| 7 | 13 | 16 | 19 | 26 |

**Supplementary Figure 1.**
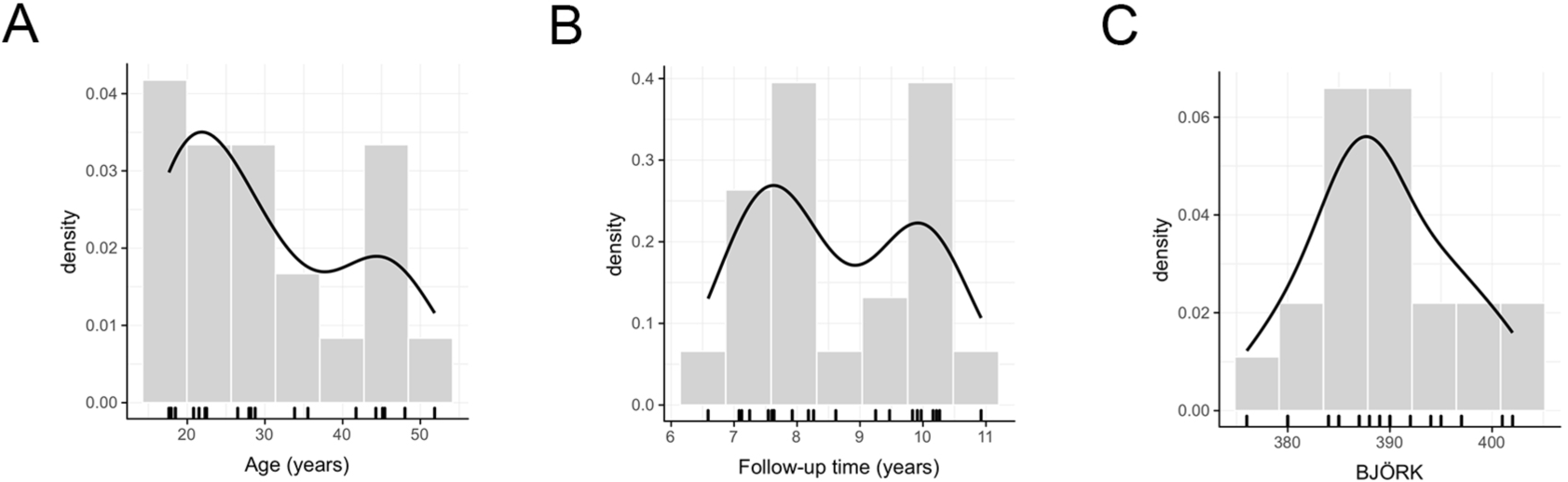
Histograms and kernel density estimates. **A** Age in years. **B** Follow-up time in years. **C** Björk’s angle sum in degrees.

